# Neural Shape Modeling Reveals Early and Progressive Femoral Bone Shape and Cartilage Thickness Changes After Anterior Cruciate Ligament Reconstruction

**DOI:** 10.1101/2025.11.16.25340352

**Authors:** Anoosha Pai S, McKenzie S. White, Marianne S. Black, Katherine A. Young, Seth L. Sherman, Constance R. Chu, Ashley Williams, Garry E. Gold, Feliks Kogan, Brian A. Hargreaves, Akshay S. Chaudhari, Anthony A. Gatti

**Affiliations:** Department of Bioengineering, Stanford University, Stanford, USA; Department of Radiology, Stanford University, Stanford, USA; Department of Orthopaedics, Stanford University, Stanford, USA; Department of Biomedical Data Science, Stanford University, Stanford, USA

**Keywords:** Post-traumatic osteoarthritis, anterior cruciate ligament reconstruction, femoral bone shape, femoral cartilage thickness, neural shape model

## Abstract

**Objectives:** To assess early and longitudinal post-traumatic osteoarthritis (POTA) changes in the femoral bone and cartilage following after anterior cruciate ligament reconstruction (ACLR).

**Methods:** CLR and contralateral knees of 17 subjects (11M, 38±11years), and 17 matched healthy controls underwent 3T-MRI at 3-weeks, 3, 9, 18, and 30 months post-ACLR. Two neural shape models (NSMs)—B-NSM (bone-only) and BC-NSM (bone+cartilage), trained on the Osteoarthritis Initiative (OAI), encoded femoral bone and cartilage shapes. The study endpoints were shape scores (B-Score, BC-Score), regional bone surface area, and cartilage thickness. Cross-sectional and longitudinal changes were assessed with linear mixed-effects models; ηp^2^ evaluated sensitivity to change. Cosine similarity compared trajectories of progressive bone and cartilage shape changes in PTOA (this data) and idiopathic-OA (OAI data).

**Results:** At 3-weeks, ACLR knee B-Scores were significantly lower (p=0.010) than contralateral knees from surgical notchplasty. Over 30-months, ACLR knee B- and BC-Scores significantly increased (p<0.001, p<0.001) compared to baseline, with no significant change in scores of other knees. Bone surface area (η_p_^2^=0.01) increased from periarticular osteophyte lipping rather than subchondral plate enlargement, with concomitant region-specific femoral cartilage thinning and thickening (η_p_^2^=0.02). B-Score (η_p_^2^=0.45) was more sensitive to longitudinal change than BC-Score (η_p_^2^=0.30). ACLR knee bone shape changes aligned with idiopathic-OA progression (cosine similarity=0.25); cartilage thickness changes did not (cosine similarity=0.09).

**Conclusion:** Early and progressive bone remodeling and spatiotemporal cartilage thickness changes were observed post-ACLR. Both B- and B+C-Scores outperformed bone surface area and cartilage thickness measures in quantifying longitudinal changes in ACLR knees. ACLR bone shape features exhibited a linear progressive pattern similar to idiopathic-OA, while the cartilage thickness changes were nonlinear. NSM-derived, reader-independent, shape metrics of the bone and cartilage may serve as early biomarkers to study PTOA in ACLRs, or as quantitative endpoints for prevention and early-intervention trials.

## INTRODUCTION

Knee osteoarthritis (OA) is a degenerative whole joint disease affecting ∼365 million people globally^1^. Post-traumatic osteoarthritis (PTOA) is a subset of OA triggered by injury or trauma to the knee joint, and constitutes 12% of all OA^2^. 23-50% of individuals with an anterior cruciate ligament (ACL) injury or ACL reconstruction (ACLR) develop PTOA within10 years^3–6^. Knee OA involves subchondral bone remodeling, characterized radiographically by the development of marginal osteophytes, narrowing of the intercondylar notch, flattening of the femoral condyles^7,8^, along with degradation of articular cartilage leading to full-thickness loss^9,10^. How these changes manifest during the onset and development of PTOA after ACLR, however, is not well understood.

Alterations in femur bone shape is proposed as a sign of future PTOA^11,12^, with changes in some shape features preceding radiographic symptoms by at least 12 months^13,14^. Prior studies on individuals with ACLR have also reported rapid enlargement of medial femur exceeding those in healthy knees in the first two years after surgery^15,16^. Quantitative MRI studies have further demonstrated greater femoral cartilage matrix degradation^17^ accompanied by thickness changes^18^ post-ACLR. Further, there is no comprehensive evaluation of overall bone and cartilage morphologies in the context of PTOA development post-ACLR. Further, PTOA is often multicompartmental, with tibiofemoral and patellofemoral compartments showing distinct changes^19^. However, the precise anatomical locations, morphological patterns, magnitudes, and co-occurrences of these early shape changes following ACLR remain unclear.

Conventional imaging metrics of knee OA (e.g. osteophyte grading, joint-space width, focal cartilage thickness, etc.) target a few isolated shape features and are often late-stage, compartment-specific and reader-dependent^20,21^. Shape modeling enables comprehensive representation and reader-agnostic quantification of continuous, 3-dimensional (3D) knee morphology associated with the disease from medical imaging data^22,23^. Shape modeling has been previously applied to the femur to encode the 3D bone geometry into a single metric of OA severity called the B-Score, which is analogous to the T-score clinically used in osteoporosis management^24^. A higher B-Score indicates a greater deviation from the healthy bone shape, reflecting a bone morphology that is more characteristic of OA. A Prior statistical shape model (SSM)-based study demonstrated that ACLR knees exhibited higher femur B-Scores than contralateral knees 2 years post-surgery^25^. However, traditional SSMs are constrained by linear assumptions and the need for dense, accurate point correspondences, which limit their ability to capture high-dimensional, nonlinear changes^23^. Neural shape models (NSM), improve upon SSMs by leveraging advanced concepts of computer graphics and deep learning to encode nonlinear and clinically relevant shape features of OA without requiring explicit point matching^26^. However, NSMs have not been applied to quantify PTOA related remodeling in ACLR individuals.

Existing shape modeling studies for OA are predominantly on the femur bone only. No study to date captures concomitant early cartilage changes that occur alongside the bone after ACLR. Hence, a combined shape score encoding 3D geometries of both bone and cartilage can provide remarkable insights on PTOA development in ACLR knees. Importantly, there are no data describing femoral bone and cartilage shape changes immediately following ACLR surgery and how these changes evolve during the early post-surgical period—a critical window when early detection can enable risk stratification and timely interventions may prevent or slow PTOA. Understanding early changes in bone and cartilage shape in PTOA can also be used to elucidate whether PTOA and idiopathic OA share the same progression patterns.

The purpose of this study was to quantify, localize, and track early and progressive PTOA–related changes in both femur and femoral cartilage after ACLR. We apply two NSMs, a bone-only (B-NSM) and a combined bone-and-cartilage (BC-NSM) to ACLR and healthy control knees to assess shape changes immediately (3 weeks) following ACLR surgery and to track them longitudinally (3, 9, 18, and 30 months). First, we compare the sensitivities of the Bone-only Score (B-Score) and the combined bone-and-cartilage score (BC-Score) to detect early and longitudinal OA-like shape changes in the ACLR cohort. Second, we spatially localize and quantify shape changes by computing regional bone surface area and regional cartilage thickness measures longitudinally. Third, we compare the overall trajectory of PTOA progression following ACLR to mean idiopathic OA progression, revealing unique joint changes in PTOA.

## METHODS

### Study Sample

The study enrolled participants from the community with a confirmed unilateral, clinically indicated ACL injury and scheduled for ACLR, along with age, sex, and BMI matched healthy controls from March 2017 to August 2024. ACLR and contralateral knees of ACLR subjects were scanned at 3-weeks (baseline), 3, 9, 18, and 30 months following ACLR. The right knee of the healthy controls was imaged at the same timepoints. All participants provided written informed consent in accordance with IRB guidelines, and the study followed the Institutional Review Board guidelines. The exclusion criteria for recruitment were BMI > 35, visible varus/valgus deformities, previous ACL tears and/or surgeries in either knee, previous injury to either knee, concurrent PCL tears, posterolateral corner injuries, displaced tibia fractures, full-thickness chondral defects, prior ligament injuries or surgeries, Kellgren-Lawrence grade > 1, and any contraindications to MRI.

### Imaging protocol

All subjects were scanned in a 3T scanner (GE Healthcare, Waukesha, WI). MRI scanning was performed on each leg unilaterally using a 16-channel receive-only flexible extremity coil (Neocoil, Pewaukee, WI) using a quantitative double-echo in steady-state (qDESS) sequence (TE_1_/TE_2_ = 6/38 ms, TR = 22 ms, flip angle = 25°, slice thickness = 1.5 mm, number of slices = 80, FOV = 16.0 cm, bandwidth = ±31.25 kHz, acquisition matrix = 384 × 320, phase acceleration: 2 x 1, total acquisition time = 9 minutes).

### Image and Shape Analysis

The femoral bone and cartilage were automatically segmented on the qDESS scans using DOSMA, a deep-learning, open-source framework for musculoskeletal MRI analysis^27^ and a previously validated automated pipeline^28^ (Supplementary Figure 1). Bone and cartilage segmentation masks were used to generate 3D surface meshes^29^ (Supplementary Figure 1). To remove variance due to laterality, all left side meshes were mirrored to represent and align with the right-side knee.

The femoral cartilage thickness was computed from the femoral cartilage surface mesh and was represented as a mean projection on to the bone surface mesh. The cartilage thickness projection at any point on the bone surface was computed as the Euclidean distance between two points of intersection of the normal vector with the cartilage mesh, where the normal vector is projected from the vertex on the bone mesh.

### Neural Shape Modeling

Two NSMs were developed from the baseline double-echo in steady-state (DESS) scans from 3,233 subjects and 6,325 knees varying across KL grades 0 to 4 from the Osteoarthritis Initiative (OAI)^23^. The B-NSM, was trained only on 3D femoral bone surface, and the BC-NSM was trained on both the bone and cartilage 3D surfaces.

Each NSM model encodes 512 shape features, which are then used to generate an “idiopathic OA progression vector”, such that its origin (Score = 0) represents the average healthy shape features (KL grade = 0 at all timepoints) and the positive direction indicates progressive OA-like changes (KL grade > 0 at all timepoints) in each of those 512 shape features (Supplementary Figure 2A). The B-Score/BC-Score is defined as the distance from the average healthy shape (Score=0) along this vector. One unit on this vector represents one standard deviation from the mean healthy knee shape. To characterize idiopathic OA shape features encoded in each NSM, we simulated synthetic OA progression along this vector. We generated reconstructions of bone surface (using B-NSM) and bone and cartilage surfaces (using BC-NSM) for various B- or BC-Scores (0, 2, 4, and 6). The bone shape and cartilage thickness changes at each score with respective to the mean healthy shape (B- or BC-Score =0) was visualized (Figure 1).

**Figure 1:**
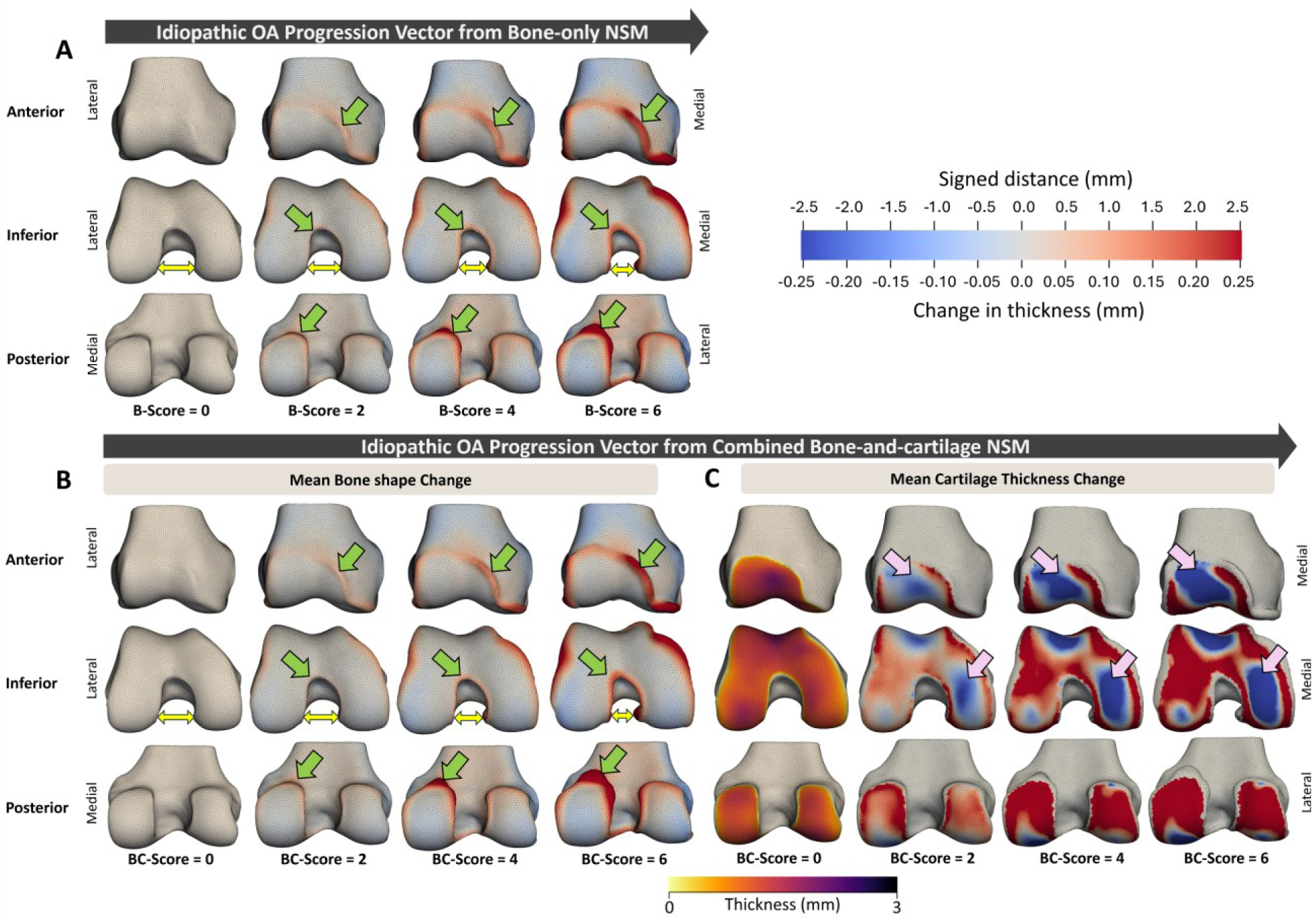
OA progression simulated using NSMs: femoral bone and cartilage-thickness changes displayed for discrete shape scores along the idiopathic OA progression vector (generated from the OAI dataset). **A)** Femoral bone shape changes from a bone-only NSM. With increasing OA severity/B-Score (left to right) the bone reveals a narrowing of the intercondylar notch (double-sided yellow arrow), widening and flattening of the condyles and osteophytes surrounding the cartilage plate (green arrows). **B)** Femoral Bone shape and **C)** femoral cartilage thickness changes from a combined bone-and- cartilage NSM. Bone shapes are similar to bone-only NSM. The femoral cartilage shows thinning in the anterior patellofemoral, and medial tibiofemoral region with OA severity (pink arrows) compared to the mean healthy (BC-Score=0). Colormap: Change in bone shape at each B-score is represented as signed-distance with respect to the mean healthy bone (B-score=0): Red (positive)=outward protrusion; beige (zero)=no change; blue (negative)=inward contraction. Cartilage thickness changes are represented as change from the mean healthy thickness map (BC-Score=0): Red (positive)=thickening; beige (zero)=no change; blue (negative)=thinning. B-Score=shape score generated from the Bone-only NSM; BC-Score=shape score generated from the combined bone-and-cartilage NSM. OA=Osteoarthritis, NSM =Neural shape model.

### Quantifying Shape Change

To visualize bone shape change, the signed-distance of each point on one mesh was computed with respect to the corresponding point on the reference mesh. For a given bone surface, the “sign” of this distance is positive for points that are outside (red color in Figure 1A and B), zero for points that are on (beige color), and negative for points that are inside the reference bone surface (blue color). Bone regions that are outside indicate enlargement while inside indicates shrinkage. To assess change in cartilage thickness between any two meshes, we computed the difference between the cartilage thickness (projected on to the bone surface) on a given mesh with respect to the reference mesh. Similar to the signed-distance, a positive value (red color) indicates thickening, zero (beige color) indicates no change, and a negative value (blue color) indicates thinning of the cartilage compared to the reference thickness (Figure 1C).

### NSM Application to ACLR and Healthy Control Cohort

#### B-Score and BC-Score

To remove the bias of subject size and position, we initially registered the bone and cartilage surface meshes from our study to a normalized atlas mesh. This atlas mesh is the average mesh across all healthy subjects (KL grade = 0) from the OAI data. We then fit the B-NSM and the BC-NSM to the meshes of each ACLR, contralateral, and healthy control knee at every timepoint to compute B-Scores and BC-Scores, respectively (Supplementary Figure 1). To evaluate shape differences immediately after surgery, we compared the B-Scores and BC-Scores between the ACLR, contralateral, and healthy control knees at the baseline visit (3 weeks post-surgery). To assess longitudinal progression, we calculated change in B- and BC-Scores at each follow-up visit with respect to the baseline visit (follow-up minus baseline).

#### Shape Changes

To visualize shape changes between knee groups and across time points, we generated a “mean shape” by averaging the shape features of all subjects in each knee group at every timepoint. Cross-sectional changes immediately post-ACLR were visualized using signed-distance of mean ACLR knee with respect to the mean contralateral knee (Figure 2)^30^.

**Figure 2:**
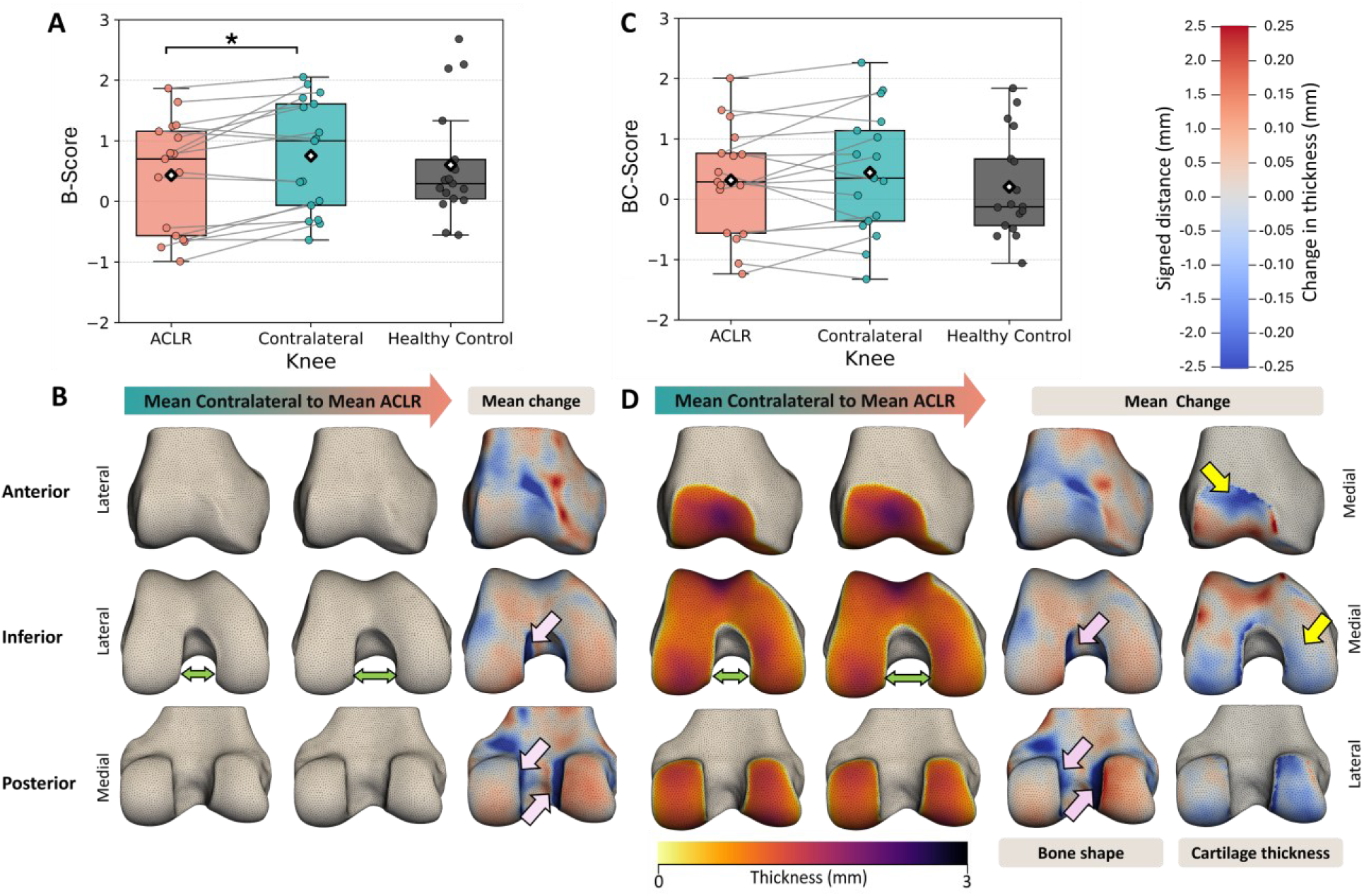
Femoral bone and cartilage thickness changes at 3-weeks following surgery. **A)** B-Score for ACLR knee was significantly lower than the contralateral knee (η_p_^2^=0.34, p=0.010), but not different from the healthy control knee (p=0.848). **B)** Visualization from the B-NSM. ACLR knees exhibit a wider intercondylar notch (∼5 mm wider, double-sided green arrows or blue regions pointed by pink arrows) compared to contralateral knee resulting from surgical notchplasty, yielding a shape less characteristic of OA (lower B-Score). **C)** The BC-Scores were not significantly different between knee-types (η_p_^2^=0.07, p=0.473). **D)** Visualization from the BC-NSM. Femoral cartilage in ACLR knees was thinner in the anterior and the medial regions (yellow arrows) compared to contralateral knees. Bone shape changes between were similar to B-NSM. Colormap: Change in ACLR bone shape is represented as signed-distance with respect to the contralateral bone at baseline visit: Red (positive)=outward protrusion; beige (zero)=no change; blue (negative)=inward contraction. Cartilage thickness changes are represented as change from the contralateral thickness map at baseline visit Red (positive)=thickening; beige (zero)=no change; blue (negative)=thinning. Effect size (η_p_^2^); small (η_p_^2^≥0.01), medium (η_p_^2^≥0.06), or large (η_p_^2^≥0.14). OA=Osteoarthritis, NSM=Neural shape model.

Longitudinal shape changes for each knee-type were visualized using signed-distance at each follow-up visit compared to those at the baseline visit (Figure 4).

#### Regional Bone Surface Area and Femoral Cartilage Thickness

To quantify localized changes in bone surface area, we defined three consistent and clinically relevant bone regions (Supplementary material, Bone Region Definitions): the subchondral region, representing bone area immediately beneath the articular cartilage (Figure 5A), the periarticular region, capturing the marginal rim near the articular cartilage edge where osteophytes typically develop (Figure 5B), and a composite region defined as the union of the subchondral and periarticular regions (Figure 5C). Each bone region was further subdivided into anatomical subregions—anterior, medial-central, medial-posterior, lateral-central, and lateral-posterior based on previously established standards^31^. For each bone region, we computed bone surface area within every anatomical subregion. We also measured the mean cartilage thickness within each anatomical subregion for ACLR, contralateral, and healthy control knees at every timepoint (Figure 5D).

### Statistical Analysis

#### Assessing Early And longitudinal Changes in B- and BC-Score After ACLR

We employed linear mixed effects models (LMEM) to examine the effect of (i) “knee-type” and “timepoint” on B- and BC-Scores at baseline visit, and longitudinally; (ii) “knee-type”, “timepoint”, and “anatomical subregion” on longitudinal regional bone surface area and cartilage thickness changes. “Knee-type” (reference: ACLR) and “subregion” (reference: lateral-central) were treated as categorical variables and “timepoint” was represented as a continuous measure indicating the number of months after ACLR surgery. Our model included fixed effects (knee-type, timepoint, and anatomical subregion), two-way (knee-type × timepoint, knee-type × anatomical subregion, timepoint × anatomical subregion) and three-way interaction effects (knee-type × timepoint × anatomical subregion), and random effects (participant).

Each LMEM was fit using the restricted maximum likelihood (REML) method employing the Broyden–Fletcher–Goldfarb–Shanno algorithm for unconstrained optimization. The p-values for the fixed effects were calculated using Satterthwaite’s method^32^. We assessed the normality of the residuals from the LMM using the Shapiro–Wilk test. When fixed or interaction effects were significant, pairwise comparisons with Tukey’s corrections were performed. Effect sizes for significant fixed and interaction effects were computed as partial eta squared (η_p_^2^); defined as small (η_p_^2^ ≥ 0.01), medium (η_p_^2^ ≥ 0.06), or large (η_p_^2^ ≥ 0.14)^33^. For our analysis, we interpreted a higher effect size as a higher sensitivity to detect change. Values with p<0.05 were considered significant. Statistical analysis was performed in R (v4.3.2), using rpy2 (v3.5.15) python module.

#### Comparing PTOA and Idiopathic OA Progression Trajectories

We first represented each knee by 512 NSM-based shape features. For each knee group, we fit a LMEM with these shape features as independent variables, timepoint as the dependent variable, and subjects as a random effect. The fixed effects of this LMEM represents “ACLR-to-PTOA progression vector”, which summarizes the direction and magnitude of average longitudinal changes in each of the 512 shape features while accounting for individual variability (Supplementary Figure 2B). To compare PTOA shape trajectory post-ACLR with idiopathic OA trajectory, we compute the cosine similarity between the ACLR-to-PTOA progression vector and the idiopathic OA progression vector (generated from the OAI)^23^. Cosine similarity was interpreted as follows: +1 denotes complete alignment in the direction of shape change, 0 denotes orthogonal (unrelated) directions, and −1 denotes diametrically opposed trajectories, indicating divergent progression pattern. Separately, we compared progression trajectories derived from the B-NSM and the BC-NSM.

## RESULTS

### Study Population

Seventeen ACLR subjects (sex= 11 male, 6 female, age= 38±11 years, height= 174±9cm, mass= 70.6±9.3kg, body mass index (BMI)= 23.3±1.6 kg/m^2^) and 17 healthy controls (sex= 11 male, 6 female, age= 38±12 years, height= 175±8cm, mass = 75.4±12.3kg, body mass index (BMI)= 24.5±2.8 kg/m^2^) were included. Out of the 34 subjects, 12 missed at least one visit: 9 (4 ACLRs, 5 controls) missed 1 of 5 visits, 1 ACLR subject missed 2 visits, and 2 ACLR subjects missed 3 visits; the remaining 22 subjects completed all 5 visits. Overall, out of a total of 255 data points [51 knees= 17 ACLR + 17 contralateral + 17 control) × 5 visits], 226 (88.63%) were successfully collected.

### Changes Immediately Following ACLR

At post-surgery baseline, ACLR knee B-Score was significantly lower than the contralateral knee (η_p_^2^=0.34, p=0.010; Figure 2A), but not significantly different from the healthy control knee (p=0.848). The BC-Scores were not significantly different between the knee-types (η_p_^2^=0.07, p=0.473; Figure 2C).

### Longitudinal And Progressive Changes Following ACLR

Longitudinally, both B- and BC-Scores showed significant knee × timepoint interaction effect. The B-Score in ACLR knees showed significantly greater increases (η_p_^2^=0.45, Figure 3A) over 30 months compared to contralateral (p<0.001) and healthy control knees (p<0.001). The change in BC-Score in ACLR knees also significantly increased with time (η_p_^2^=0.30, Figure 3B), compared to the contralateral (p<0.001) and the healthy control knees (p<0.001).

**Figure 3:**
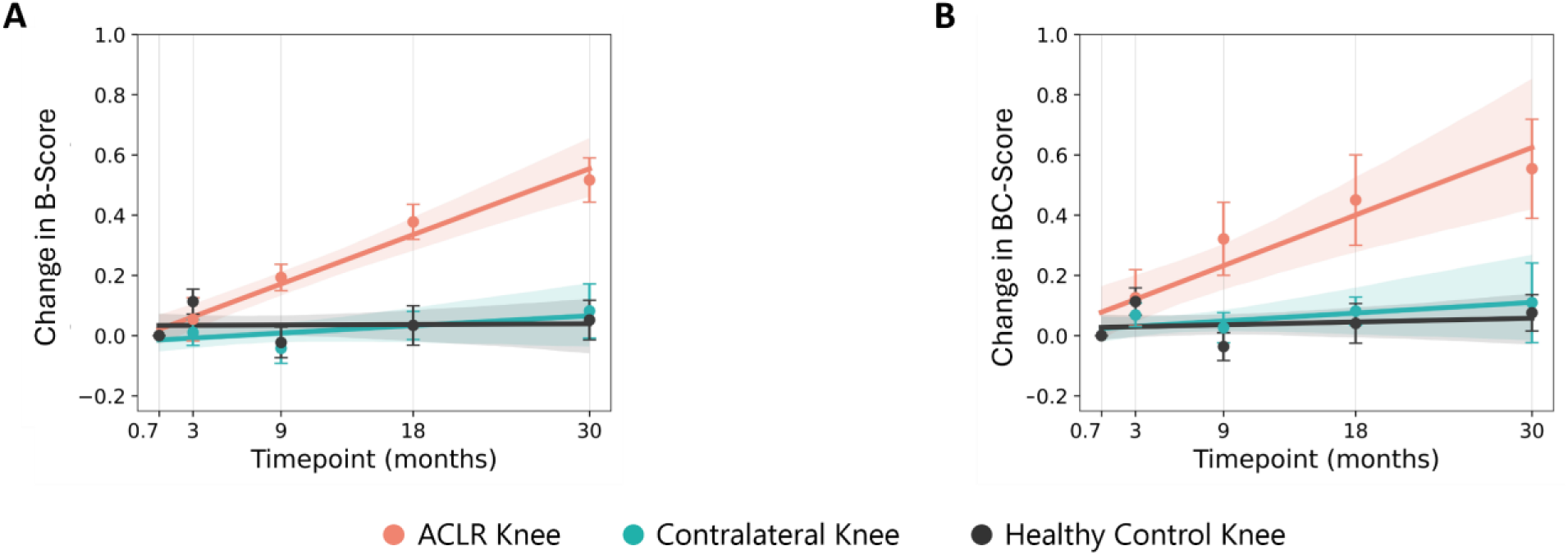
Longitudinal change in B-Score progression with respect to baseline (3-weeksfollowing surgery). The dot and the error bars represent the raw mean and the standard errors at each timepoint. The shaded region represents the 90% CI of the linear regression fit. **A)** B-Score: shape score from the bone-only NSM. The ACLR knees showed significantly greater increases (η_p_^2^=0.45) in B-Score over 30 months compared to the contralateral (p<0.001) and healthy control knees (p<0.001). **B)** The BC-Score: shape score from the combined bone-and-cartilage NSM. The ACLR knees showed significantly greater increases in BC-Score with time (η_p_^2^=0.30), compared to the contralateral (p<0.001) and the healthy control knees (p<0.001). The B-Score demonstrated higher sensitivity (effect size) and lower between-subject variability (smaller confidence band) than the BC-Score in detecting progressive PTOA-related changes in the ACLR knees. Effect size (η_p_^2^); small (η_p_^2^≥0.01), medium (η_p_^2^≥0.06), or large (η_p_^2^≥0.14). NSM = Neural shape model.

### Bone Surface Area and Cartilage Thickness Changes Following ACLR

In all anatomical subregions, ACLR knees exhibited greater increases in composite (p<0.006, η_p_^2^=0.01, 1.7%) and periarticular (p<0.026, η_p_^2^=0.02, 5.34%) bone surface areas over time compared to contralateral and healthy control knees (Figure 5A, B, and C). The rate of increase (slope) in the composite (η_p_^2^=0.05, Figure 5C) and periarticular (η_p_^2^=0.06, Figure ^5^B) bone surface area for the ACLR knees was significantly higher in the anterior (composite: p<0.001, periarticular: p<0.001), medial-posterior (composite: p=0.023, periarticular: p=0.002), and lateral-posterior (composite: p=0.008, periarticular: p=0.001) regions compared to the same anatomical subregions in contralateral and healthy control knees during 3 to 30 months post-ACLR. Subchondral bone surface area showed no significant (p=0.154) longitudinal change in any anatomical subregion (Figure 5A).

The regional cartilage thickness showed significant knee-type × timepoint × anatomical subregion interaction effects (η_p_^2^=0.02, p=0.002, Figure 5D). While rate of change in cartilage thickness (slope) over time showed a significant decrease in the anterior region (p=0.006), it exhibited significant increase in medial-central (p<0.001), medial-posterior (p=0.016), and lateral-central (p=0.042) regions for the ACLR knees when compared to the slopes of contralateral and healthy control knees in the same regions.

### PTOA vs Idiopathic OA Progression

The cosine similarity between the ACLR-to-PTOA progression vector and the idiopathic OA progression vector using bone-only features was 0.25 for ACLR knees, –0.03 for contralateral knees, and –0.09 for control knees. When using combined bone-and-cartilage features, the cosine similarity was 0.09 for ACLR knees, 0.04 for contralateral knees, and –0.04 for control knees compared to the idiopathic OA trajectory.

## DISCUSSION

In this study, we used NSMs to effectively characterize early and longitudinal shape changes in ACLR femoral bone and cartilage. At the 3 weeks post-surgery baseline, ACLR knees displayed lower B-Score compared to the contralateral knee likely due to surgical widening of the intercondylar notch. Over 30 months post-surgery, ACLR knees exhibited substantial increases in both B- and BC-Scores, compared to contralateral and healthy control knees indicating OA-like progressive changes in both bone and cartilage morphology. Specifically, ACLR knees exhibited steep increases in bone surface area primarily driven by periarticular osteophyte lipping rather than subchondral plate enlargement along with concomitant, region-specific thinning and thickening of femoral cartilage. While both B- and BC-Scores were more sensitive than both bone surface area and cartilage thickness measures in detecting longitudinal change, the B-NSM was more sensitive in detecting progressive OA-like features compared to the BC-NSM. Our results indicate that PTOA bone shape changes in ACLR knees exhibit linear progressive pattern similar to the idiopathic OA. In contrast, the cartilage thickness changes are more heterogenous and nonlinear in nature.

### Post-Surgical Baselines

At baseline, a lower B-Score in ACLR knees indicates that the surgical knee had a less OA-like bone shape compared to the contralateral knee. Visualizations from the B-NSM revealed that the ACLR knees had a wider (∼5mm) intercondylar notch than their contralateral knee, which resulted from notchplasty confirmed on surgical notes (Figure 2B). A notchplasty may be performed during an ACLR, to aid in surgical visualization, to treat limited knee extension in patients, and to prevent ACL impingmenet^34^. Since an idiopathic OA femur typically has flatter condyles leading to a narrower intercondylar notch^35,36^ (Figure 1 A and B), the surgically altered ACLR bone geometry appeared to be less characteristic of OA, yielding a lower B-Score. On the other hand, the BC-Scores were not significantly different between the knees. Reconstructions from the BC-NSM also revealed that the ACLR bone had a wider notch compared to their contralateral knee (Figure 2D). However, the femoral cartilage in ACLR knees was thinner in the anterior and the medial regions (Figure 2D) when compared to that of the contralateral knees, which is consistent with greater OA disease burden (Figure 1C). BC-Score is a metric that encodes OA-like shape features from both bone and cartilage. Consequently, the combination of healthy-like bone shape and OA-like cartilage thickness in ACLR knees resulted in BC-Scores similar to those of contralateral and healthy control knees at baseline. Our results suggest that postoperative knee shape obtained at the earliest timepoint after surgery, which is inclusive of surgery-related changes, is needed to enable longitudinal PTOA monitoring after ACLR.

### Bone Shape Changes

Longitudinally, the B-Score of ACLR knees significantly increased over 30 months compared to the post-surgery baseline, while the contralateral and control knee B-Scores showed no change with time. Particularly, ACLR knees showed modest yet larger increases in composite bone surface area compared to the other knees, indicating bone remodeling post-surgery, a finding in-line with previous work^15^. However, steep rise in ACLR composite bone surface area over 30 months is primarily driven by increased periarticular area rather than flattening of subchondral plate (Figure 5)^37–39^. Substantial periarticular bone surface area increase suggests early and progressive osteophyte lipping, particularly prominent in the trochlea, medial-posterior and the lateral-posterior condyles (Figure 4A). Our findings are in line with previous research that demonstrated a direct correlation between regional osteophyte volume and OA severity^40,41^. These localized early osteophytes could likely be attributed to patellofemoral disruption due to grafts, lateral surgical tunnel-related remodeling, or broader changes in the mechanical environment of the ACLR joint^16,42,43^. Overall, the observed bone shape trajectory in ACLR knees align more closely (high cosine similarity) with idiopathic OA progression indicating early onset of OA-like bone remodeling following surgery.

**Figure 4:**
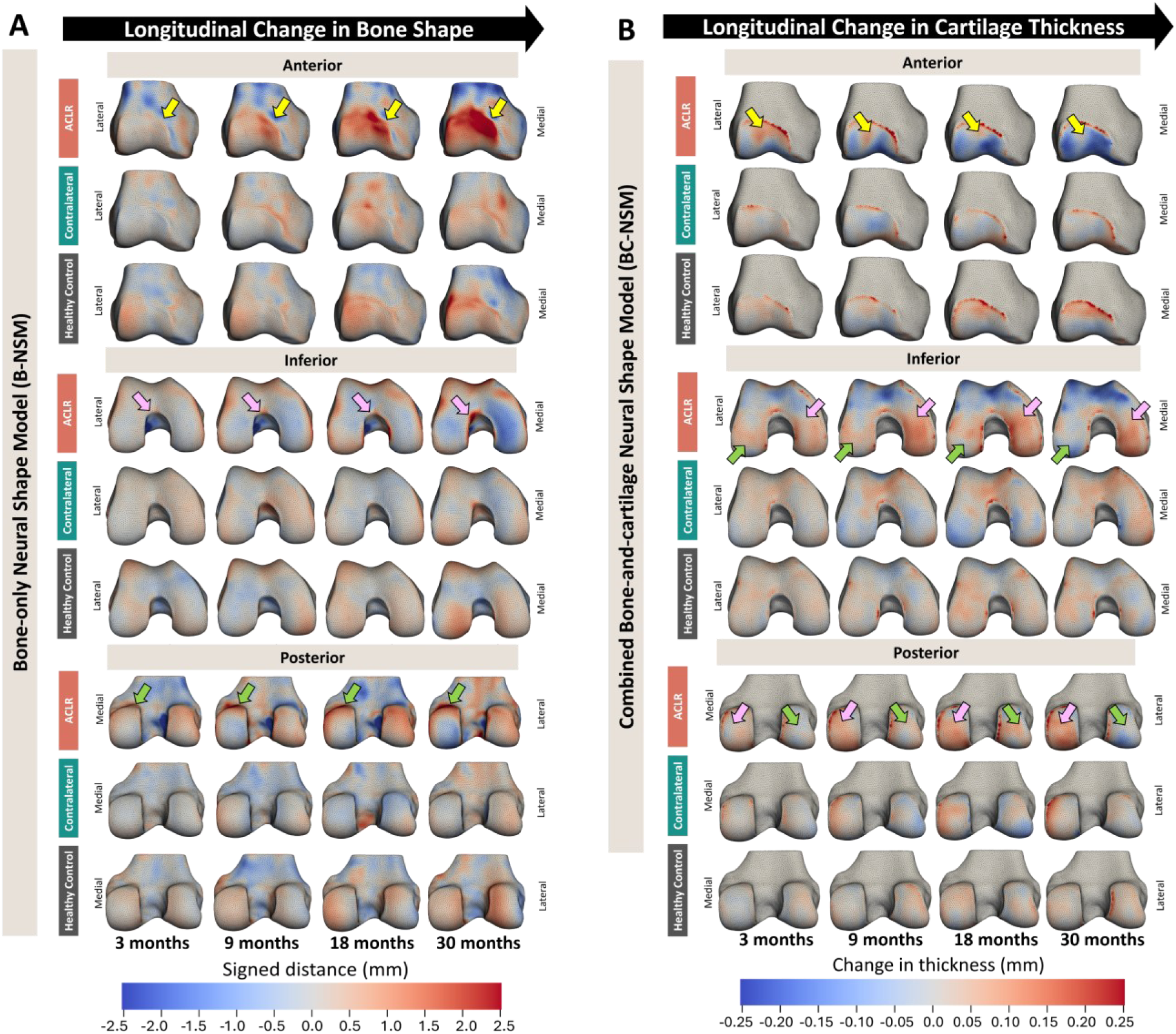
NSM-based visualizations of longitudinal shape changes in ACLR, contralateral, and healthy control knees at 3, 9, 18, and 30 months (left to right) with respect to the baseline (3-weeks following surgery). **A)** Femoral bone shape. ACLR bones show early osteophytes in the medial trochlea (yellow arrows), intercondylar notch (pink arrows), and medial-posterior condyle (green arrows)—bone shape changes that align with idiopathic OA progression. The contralateral and healthy control knees show minimal, random, and spatially diffused changes indicating no consistent change. **B)** Femoral cartilage thickness. ACLR knee displayed varying rates of thickening and thinning across cartilage subregions that did not parallel the idiopathic OA trajectory. The anterior-region showed initial thickening (3-months), followed by persistent thinning (∼0.18 mm, yellow arrows), the lateral-posterior region thickened (first 18-months) and then thinned (∼0.25mm, green arrows), whereas the medial-region underwent continuous thickening (∼0.17mm, pink arrows). This pattern, early cartilage thickening preceding eventual thinning, mirrors OA progression. Colormap: Change in bone shape is represented as signed-distance from bone shape at baseline visit: Red (positive)=outward protrusion; beige (zero)=no change; blue (negative)=inward contraction. Cartilage thickness changes are represented as change from baseline visit: Red (positive)=thickening; beige (zero)=no change; blue (negative)=thinning. OA=Osteoarthritis, NSM=Neural shape model.

**Figure 5:**
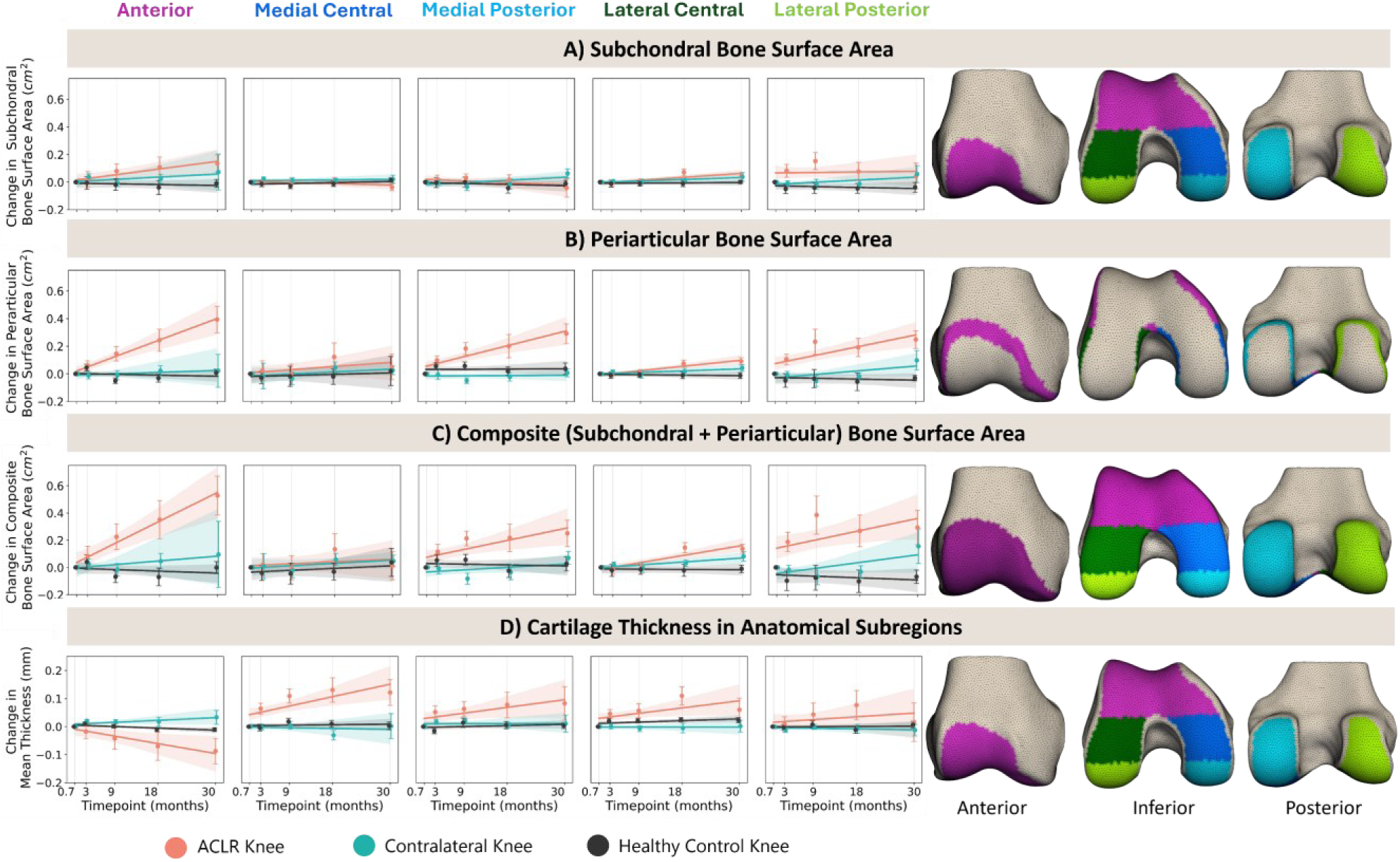
Femoral bone surface area changes in ACLR, contralateral, and healthy control knees at 3, 9, 18, and 30 months following surgery. **A)** Subchondral, **B)** Periarticular, and **C)** Composite (subchondral+periarticular) bone regions. Each bone region is subdivided into five anatomical subregions—anterior, medial central, medial posterior, lateral central, and lateral posterior. Longitudinally, ACLR knees exhibited greater increases in composite (p<0.006, η_p_^2^=0.01, 1.7%) and periarticular (p<0.026, η_p_^2^=0.02, 5.34%) bone surface areas compared to other knees. The rate of increase (slope) in the composite (η_p_^2^=0.05) and periarticular (η_p_^2^=0.06) surface area for the ACLR knees was significantly higher in the anterior (composite: p<0.001, periarticular: p<0.001), medial-posterior (composite: p=0.023, periarticular: p=0.002), and lateral-posterior (composite: p=0.008, periarticular: p=0.001) regions compared to other knees. **D)** Femoral cartilage mean thickness in each anatomic subregion. Rate of change in thickness (slope) significantly decreased in the anterior region (p=0.006), increased in medial-central (p<0.001), medial-posterior (p=0.016), and lateral-central (p=0.042) regions for the ACLR knees when compared to the slopes of other knees. The dot and the error bars represent raw mean and the standard errors at each timepoint; shaded regions represent 90% confidence-interval of the linear regression fit. Effect size (η_p_^2^); small (η_p_^2^≥0.01), medium (η_p_^2^≥0.06), or large (η_p_^2^≥0.14).

### Cartilage Thickness Changes

The ACLR knee BC-Score increased over 30 months compared to the other knees. While the bone indicated OA-like shape changes, cartilage displayed a more dynamic, nonlinear pattern that did not parallel the idiopathic OA trajectory (smaller cosine similarity)^44^. While the anterior region showed initial thickening in the first 3 months, followed by persistent thinning, the lateral-posterior region thickened during the first 18 months and then thinned, and the medial region underwent continuous thickening throughout the study period (Figure 4B and Figure **5**D). Overall, the rates of thickening and thinning varied across different anatomic subregions in our ACLR cohort, consistent with a previous study^45^. Specifically, the patellofemoral region showed accelerated degradation compared to the tibiofemoral regions, with substantial differences between subregions emerging by 18 months post-surgery. This pattern, characterized by early cartilage thickening preceding eventual thinning, mirrors previous studies documenting cartilage thickening during the early stages of OA, followed by eventual complete loss of articular cartilage in advanced OA^46,47^. Specifically, ACLR knees exhibited thinning in the anterior patellofemoral (∼0.25mm) and lateral-posterior femoral cartilage (∼0.15mm) and thickening in the medial-central region (∼0.17mm). These observations are consistent with studies that showed 1 year after ACL injury, 45% of individuals develop partial-thickness cartilage lesions in the patellofemoral joint, 29% in the medial tibiofemoral joint, and 26% in the lateral tibiofemoral joint^48,49^. These findings highlight the multicompartmental nature of PTOA and underscores the need for concurrent investigation of tibiofemoral and patellofemoral OA pathophysiology in ACLR.

### NSM Sensitivity in Capturing Progressive Shape Changes

In general, both the B- and BC-scores had greater sensitivity (larger effect sizes) to detect progressive shape changes than traditional measures such as bone surface area, and cartilage thickness. Particularly, the B-Score demonstrated higher sensitivity and lower between-subject variability (smaller confidence band, Figure 3) than the BC-Score in detecting longitudinal OA-like shape changes after ACLR. This likely reflects the more linear progression of bone shape that the B-Score captures well, whereas cartilage morphology changes nonlinearly with region-specific, concurrent thickening and thinning leading to increased variability. However, this heterogeneity in cartilage trajectories may have prognostic value in PTOA risk stratification. Future work should investigate how combined changes in bone and cartilage map to pathological modifications leading to PTOA. Our findings also suggest exploration of a PTOA-specific BC-Score to better represent the unique features.

### Limitations

Our study has several limitations. First, the sample size was modest and derived from a single center, possibly limiting generalizability. Second, because the NSMs were trained using OAI KL grade labels, which are coarse, compartment-agnostic markers validated mainly for tibiofemoral OA^50^, patellofemoral and early-stage OA features may not be adequately captured^50^. Third, despite the NSM being nonlinear, the idiopathic OA progression vector is built using linear interpolation, which mostly denotes progressive shape trajectories and is unlikely to capture non-linear changes such as initial thickening followed by thinning of cartilage. Finally, while our follow-up period was relatively long, longer-term studies are needed to fully understand predictive capability of these NSM-based metrics in future verifiable PTOA diagnosis.

## CONCLUSION

We characterized, localized, and quantified femoral bone shape and cartilage thickness changes in ACLR, contralateral and healthy control knees from 3 weeks to 30 months after ACLR surgery using a bone-only NSM and a combined bone-and-cartilage NSM. Accounting for surgically induced shape changes enabled early detection of OA-like features post-ACLR. Over 30-months post-surgery, bone shape changes in ACLR knees progress linearly and similar to idiopathic OA. In contrast, cartilage alterations follow a nonlinear and spatiotemporally varying trajectory. These observations highlight the need to concurrently assess multiple tissue compartments to fully understand and monitor the development of PTOA after ACLR. Our results support the use of NSM-derived, reader-independent, shape metrics to serve as early biomarkers to study PTOA, or as quantitative endpoints for prevention and early-intervention trials.

## Data Availability

All data produced in the present study are available upon reasonable request to the authors.

## LIST OF ABBREVIATIONS

ACLR: Anterior Cruciate Ligament Reconstruction
KL: Kellgren-Lawrence
MRI: Magnetic Resonance Imaging
NSM: Neural Shape Model
OA: Osteoarthritis
PTOA: Post-traumatic osteoarthritis
SSM: Statistical Shape Model

## DECLARATIONS

### Ethics Approval and Consent to Participate

Ethical approval for this study was obtained from Stanford University Institution Review Board (IRB 1, eProtocol #25265)

### Availability of Data and Materials

The datasets used and/or analyzed during the current study are available from the corresponding author on reasonable request.

### Authors’ Contributions

APS, FK, BAH, ASC, and AAG participated in the conception and design of the study. FK, MB, BAH, ASC were involved in imaging protocol design and development. KAY, SLS and CRC were responsible for subject recruitment. APS, MSB, KAY, FK, ASC were responsible for data collection. APS, MSW, and AAG were involved in data analysis. The interpretation of results involved contributions from APS, AW, SLS, CRC, FK, ASC, and AAG. FK, GEG, BAH, ASC provided funding for this work. All authors have reviewed and approved the final version of this manuscript and concur with the sequence in which their names are presented.

### Use of Generative-AI and AI-assisted Technologies in Manuscript Preparation

During the preparation of this work the author(s) used ChatGPT 5.0 to enhance readability and improve language after initial drafting. After using this tool/service, the author(s) reviewed and edited the content as needed and take(s) full responsibility for the content of the publication.

## SUPPLEMENTARY MATERIAL

### Bone Region Definitions

To quantify localized changes in bone surface area, we identified consistent and clinically relevant bone regions on an atlas representing average geometry of healthy femurs. This atlas was created by fitting the previously trained BC-NSM to 505 knees from the OAI dataset with no structural damage (MRI Osteoarthritis Knee Score for cartilage morphology=0 using combined assessments of cartilage lesion size and full-thickness cartilage loss). For each point on this atlas, we computed the percentage of these healthy knees having overlying femoral cartilage at that location. We thresholded this percentage to define three femoral bone regions—Subchondral (points with >95% of the healthy knees having femoral cartilage, representing bone area immediately beneath the articular cartilage, Figure 5A), periarticular (femoral cartilage-covered points in >0% but <95% of the healthy knees, capturing the marginal rim near the articular cartilage edge where osteophytes typically develop, Figure 5B), and composite (the union of the subchondral and periarticular regions including any area of cartilage coverage across the healthy knees, denoting the entire bone–cartilage interface including the outer margin, Figure 5C).

**Supplementary Figure 1:**
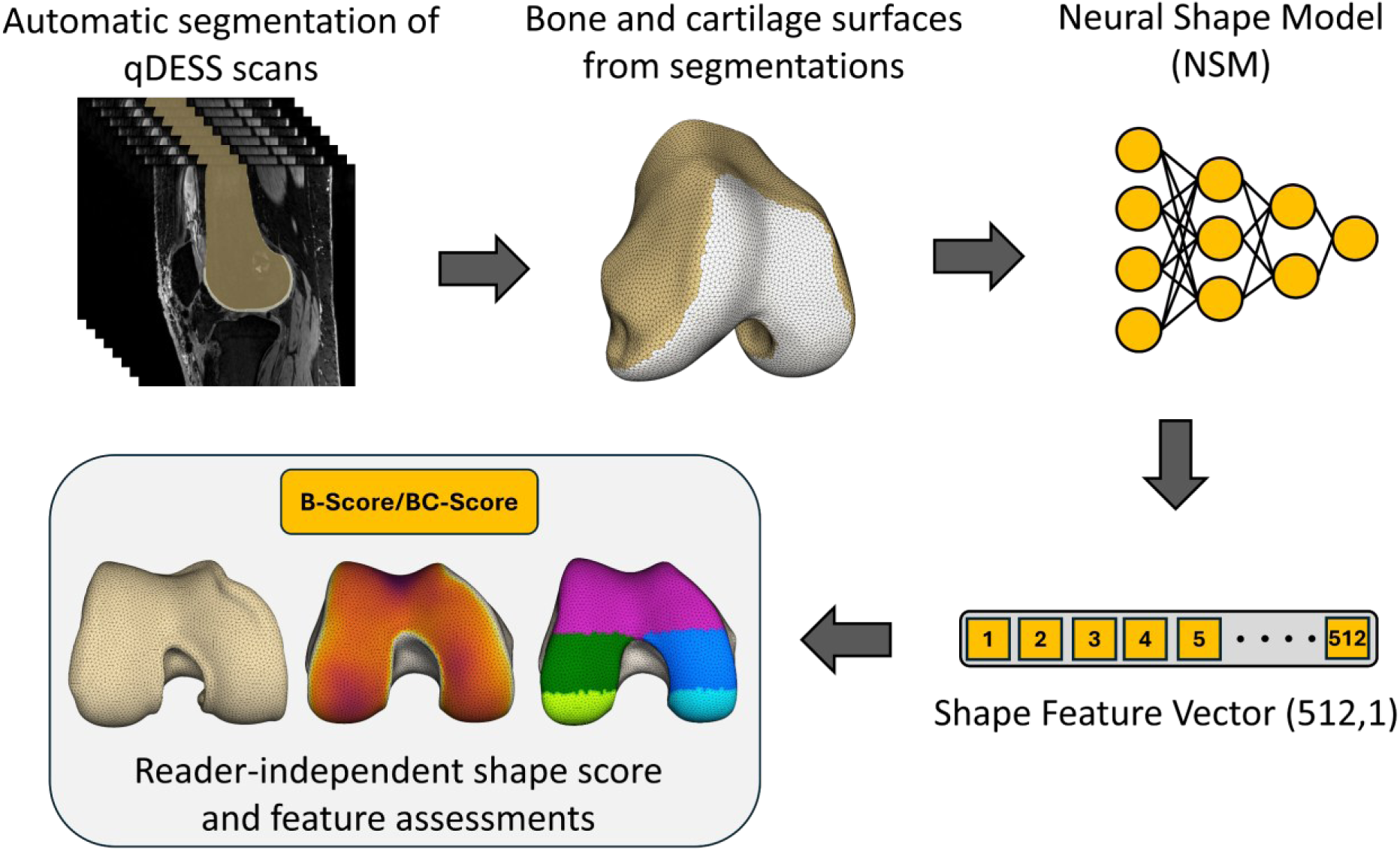
Schematic representation of the workflow for generating a reader-independent shape score and feature assessments for knee using a neural shape model. The femoral bone and the cartilage is automatically segmented on qDESS scans. Three-dimensional surfaces of the femoral bone and cartilage are generated. A neural shape model, previously trained on the OAI data, is fit to the normalized surfaces to produce a latent representation [feature vector of size (512, 1)]. This latent feature vector is then used to compute reader-independent shape scores (B-Score or BC-Score) and perform quantitative assessments (e.g., osteophyte surface area, cartilage thickness, and compartment-specific metrics). OAI=Osteoarthritis initiative.

**Supplementary Figure 2:**
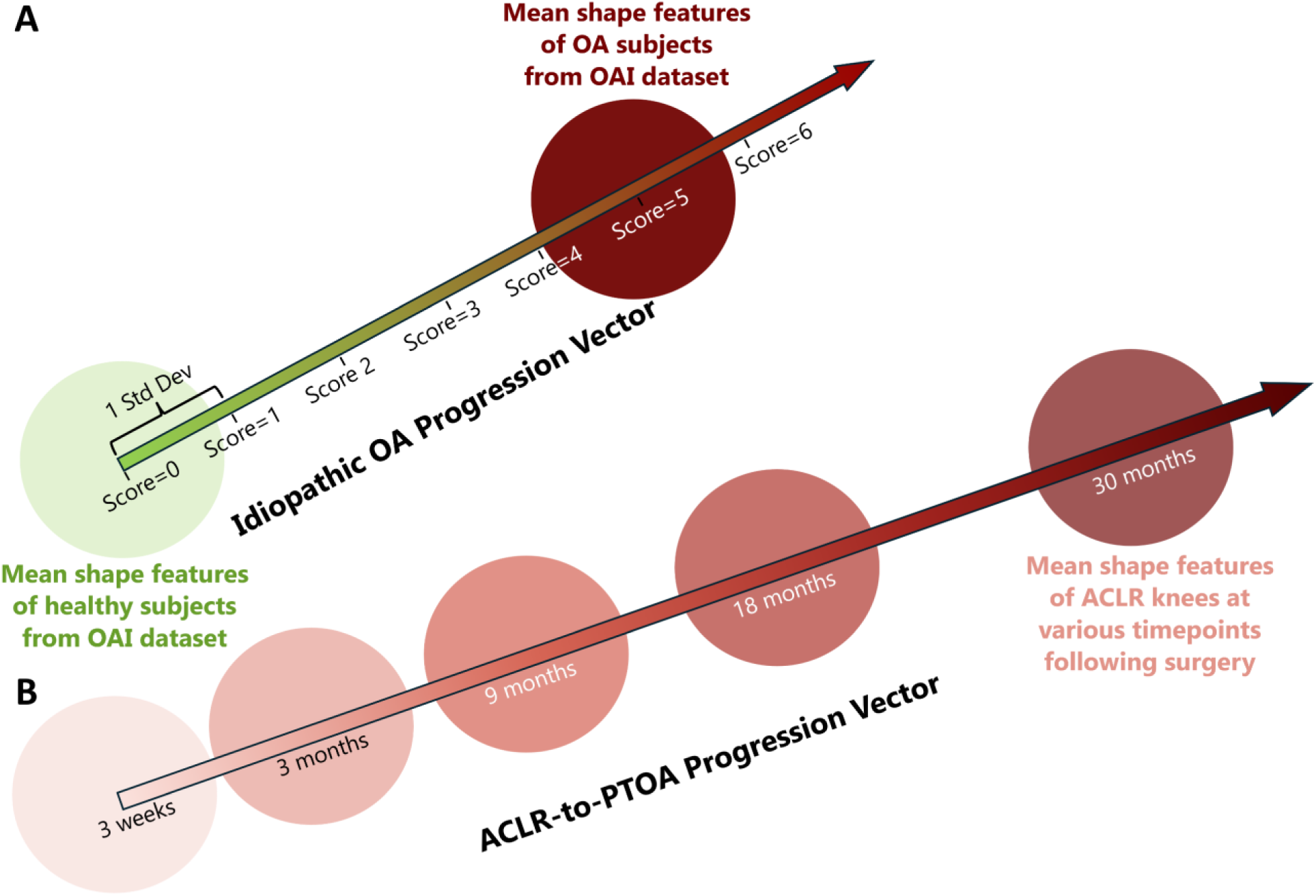
Illustration depicting progression vectors based on NSM-derived 512 shape features. **A)** Idiopathic OA progression vector defined as a vector with origin at the average healthy shape features and the positive direction indicating progressive OA-like changes in each shape features – derived from the OAI data. **B)** ACLR-to-PTOA progression vector defined as a vector with origin at the average shape features of ACLR knees at 3-weeks post-surgery, and positive direction along the mean shape features at subsequent timepoints (3, 9, 18, and 30 months). OAI=Osteoarthritis initiative.

