## Supplementary Material for "Neural Shape Modeling Reveals Early and Progressive Femoral Bone Shape and Cartilage Thickness Changes After Anterior Cruciate Ligament Reconstruction"

### Bone Region Definitions

To quantify localized changes in bone surface area, we identified consistent and clinically relevant bone regions on an atlas representing average geometry of healthy femurs. This atlas was created by fitting the previously trained BC-NSM to 505 knees from the OAI dataset with no structural damage (MRI Osteoarthritis Knee Score for cartilage morphology=0 using combined assessments of cartilage lesion size and full-thickness cartilage loss). For each point on this atlas, we computed the percentage of these healthy knees having overlying femoral cartilage at that location. We thresholded this percentage to define three femoral bone regions—Subchondral (points with >95% of the healthy knees having femoral cartilage, representing bone area immediately beneath the articular cartilage, Figure 5A), periarticular (femoral cartilage-covered points in >0% but <95% of the healthy knees, capturing the marginal rim near the articular cartilage edge where osteophytes typically develop, Figure 5B), and composite (the union of the subchondral and periarticular regions including any area of cartilage coverage across the healthy knees, denoting the entire bone–cartilage interface including the outer margin, Figure 5C).

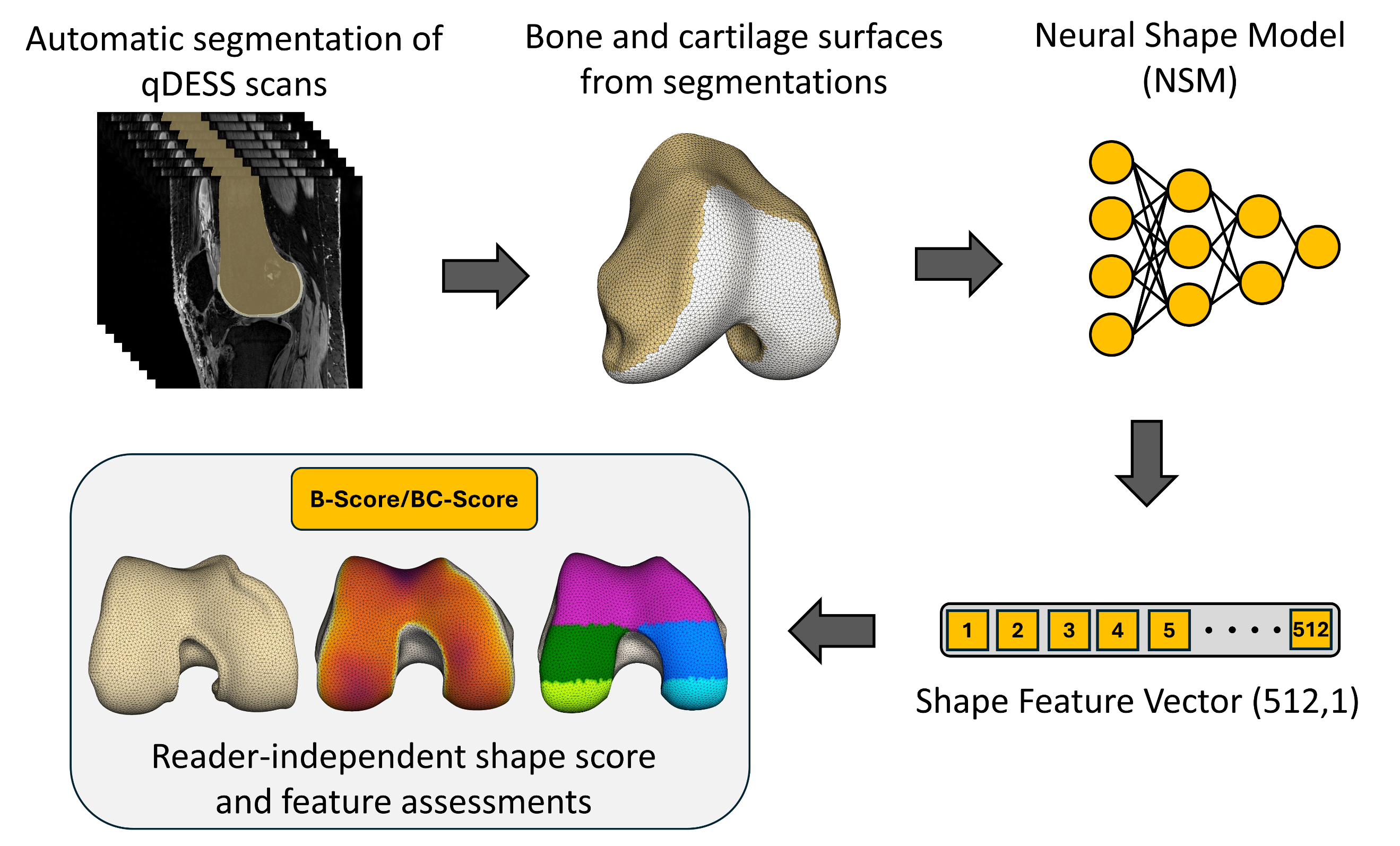

**Supplementary Figure 1:** Schematic representation of the workflow for generating a reader-independent shape score and feature assessments for knee using a neural shape model. The femoral bone and the cartilage is automatically segmented on qDESS scans. Three-dimensional surfaces of the femoral bone and cartilage are generated. A neural shape model, previously trained on the OAI data, is fit to the normalized surfaces to produce a latent representation [feature vector of size (512, 1)]. This latent feature vector is then used to compute reader-independent shape scores (B-Score or BC-Score) and perform quantitative assessments (e.g., osteophyte surface area, cartilage thickness, and compartment-specific metrics). OAI=Osteoarthritis initiative.

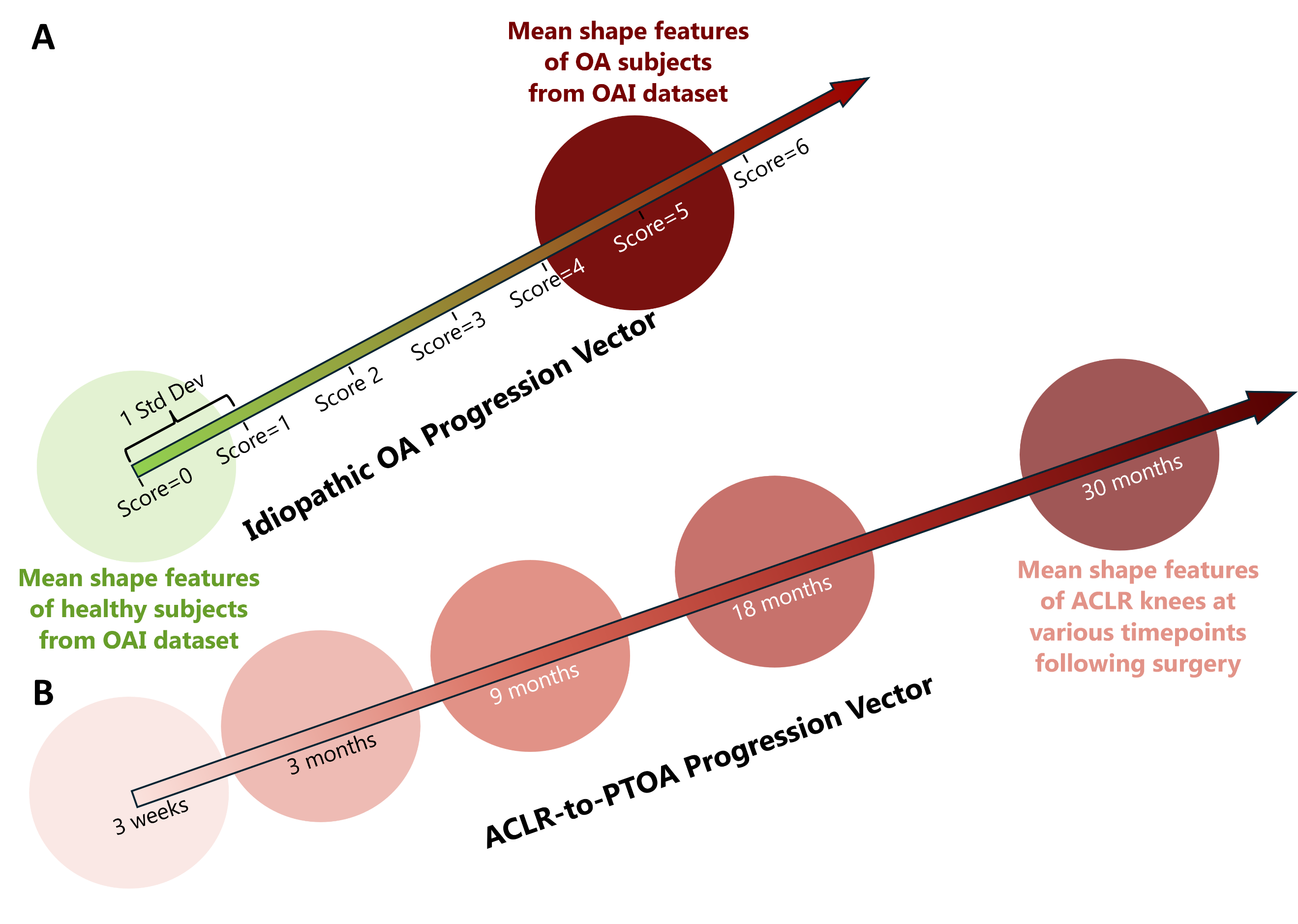

**Supplementary Figure 2:** Illustration depicting progression vectors based on NSM-derived 512 shape features. **A)** Idiopathic OA progression vector defined as a vector with origin at the average healthy shape features and the positive direction indicating progressive OA-like changes in each shape features – derived from the OAI data. **B)** ALCR-to-PTOA progression vector defined as a vector with origin at the average shape features of ACLR knees at 3-weeks post-surgery, and positive direction along the mean shape features at subsequent timepoints (3, 9, 18, and 30 months). OAI=Osteoarthritis initiative.
